# Fingolimod for treatment and or prevention of chemotherapy-induced peripheral neuropathy in humans?

**DOI:** 10.1101/2022.09.08.22279733

**Authors:** Charles L. Loprinzi, Beth Harlos, Nathan Staff, David Zahrieh

## Abstract

**Purpose:** Substantial preclinical data support that fingolimod, a drug that dysregulates sphingolipid metabolism, could both prevent chemotherapy-induced neuropathy (CIPN) and treat established chemotherapy-induced neuropathy.

**Methods:** Two pilot-phase clinical trials were developed to evaluate whether fingolimod could 1) inhibit the development of paclitaxel-induced peripheral neuropathy and 2) diminish established CIPN in humans.

**Results:** Unfortunately, both clinical trials were not able to fulfill planned accrual goals. One of the major accrual problems was the requirement for patients to be in the clinic for 8 hours following their first oral dose of fingolimod. The prevention of paclitaxel CIPN trial accrued 2 patients, one of whom did very well without any significant CIPN while the other patient developed CIPN that led to early paclitaxel dose cessation. The treatment of established CIPN trial accrued 7 of 10 planned patients. The results from the 6 patients who started daily treatment with fingolimod did not suggest that it substantially decreased CIPN over what might have been expected from a placebo effect.

**Conclusion:** Fingolimod is a clinically difficult treatment for patients to start and the limited clinical data from these protocols did not support that it was able to prevent CIPN or treat established CIPN.

## Background

Chemotherapy induced peripheral neuropathy (CIPN) is a major clinical problem that limits the utility of planned chemotherapy and can cause long-term toxicity for many patients, despite stopping neurotoxic chemotherapy. Limited options are available for treating established CIPN and there are no established means for preventing this problem, other than limiting neurotoxic chemotherapy administration.

In 2017, there was an NCI-sponsored Clinical Trials Planning Meeting designed to look at the most exciting areas for bringing forward clinical trials for chemotherapy-induced peripheral neuropathy. At that meeting, basic scientists presented their data and then these basic scientists and attending oncology clinicians/researchers met to discuss and determine which projects looked most promising to bring forward into clinical trials. Based on basic science data presented by Dr. Daniela Salvemini, fingolimod was chosen as an exciting drug to study in clinical trials.

This led to the development of two pilot clinical trials, one designed to evaluate fingolimod as an agent to prevent paclitaxel-associated CIPN and another to treat paclitaxel-associated established CIPN.

It was work in Dr. Salvemini’s lab over several years that led to the discovery that a number of chemotherapeutic agents in distinct classes and with distinct anticancer mechanisms of action (i.e., taxanes, platinum-based agents and proteasome inhibitors) drive the development of CIPN by dysregulating sphingolipid metabolism, leading to increased formation of sphingosine-1-phosphate (S1P) (1,2). They noted that the effects of S1P are mediated through activation of sphingosine-1-phosphate receptor 1 (S1PR1). Accordingly, they characterized S1PR1 antagonists, such as fingolimod, as novel non-narcotic analgesics for the treatment of chronic neuropathic pain states. This was compelling because fingolimod (3) had been FDA approved since 2010 with a reasonable safety profile (4). Salvemini and others had reported that CIPN is accompanied by significant (20-25%) partial degeneration of intra-epidermal nerve fibers (IENFs; afferent nerves’ sensory terminal arbors) (5-9) and abnormal spontaneous discharge in both A-fiber and C-fiber afferents (9-14). In rats with established bortezomib-induced peripheral neuropathy, the numbers of sural fibers with spontaneous discharge were reduced when oral fingolimod was given with bortezomib. The spontaneous discharge was felt to play a role in allodynia and hyperalgesia because onset of degeneration and peak loss of intraepidermal nerve fibers (IENF) corresponds well with pain symptoms (9,10). In animal models of CIPN, induced by paclitaxel (1), oxaliplatin (1) or bortezomib (2), oral administration of fingolimod blocked the development of, and reversed established, mechano-allodynia and mechano-hyperalgesia in a dose-dependent manner, without inducing overt leukopenia. Similar beneficial effects were observed with other S1PR1 functional antagonists, such as ozanimod and ponesimod, that were in advanced clinical trials for other indications (3,15,16). Moreover, extended treatment of fingolimod or other S1PR1 antagonists did not induce tolerance to their analgesic effects, suggesting that one could administer the drug for a long period of time and retain analgesic activity. Of note, Salvemini et al. had reported that the effects of fingolimod were not attenuated by naloxone, (1) suggesting that its effects were not dependent on engaging endogenous opioid systems.

These data led to the development of two pilot clinical trials, one to evaluate the ability of fingolimod to prevent paclitaxel-induced neuropathy and the other to evaluate whether fingolimod would be useful for treating established CIPN, in patients who had completed their neurotoxic chemotherapy.

## Methods

### Prevention of Paclitaxel-induced Neuropathy Trial

The primary goal of the prevention trial was to obtain preliminary data to support whether fingolimod would prevent CIPN in patients receiving weekly adjuvant/neoadjuvant paclitaxel therapy along with understanding the tolerability of this treatment approach in patients. The hypothesis was that we would see evidence of a marked reduction in CIPN symptoms in these patients, as measured by the EORTC-QLQ-CIPN20, compared to historical controls from three similarly conducted clinical trials. Additionally, we wanted to obtain pilot data regarding the possible relative toxicities related to fingolimod therapy in this study situation.

Eligible patients were adults scheduled to receive paclitaxel at a dose of 80 mg/m2 given every week for 12 weeks for treating breast cancer. Protocol patients needed to have a life expectancy of 6 months or more and a good performance status. They could not have had previous exposure to paclitaxel or any other neurotoxic chemotherapy. They could not have had a diagnosis of diabetic or other peripheral neuropathy nor a family history of a genetic/familial neuropathy.

At baseline, patients completed the EORTC-QLQ-CIPN20 instrument, which was then repeated prior to each subsequent paclitaxel weekly dose, at monthly intervals following the last dose of paclitaxel, and at two additional 6 month follow up visits, for a total of 18 months. Patients completed additional questionnaires at baseline and weekly to collect information regarding potential drug toxicities.

The day prior to starting their first dose of paclitaxel, patients received 0.5 mg of fingolimod; this same dose was scheduled to be repeated on the day of paclitaxel and one day after. This same fingolimod dose/schedule was to be used with each subsequent dose of paclitaxel.

With the first dose of fingolimod, patients were observed in the clinic for six hours for signs and or symptoms of bradycardia and had a post treatment EKG at 6 hours, a package label recommendation with this drug as it commonly causes temporary bradycardia that usually resolves over a few to several hours.

### Treatment of Established Neuropathy Trial

The goals for the treatment protocol were to investigate, in a pilot study, whether fingolimod markedly improved symptoms in patients with established CIPN and to evaluate the tolerability of fingolimod in these treated patients.

For the treatment trial, patients must have completed previously administered neurotoxic chemotherapy at least 3 months prior to registration. Additionally, they must have had symptoms of CIPN (tingling, numbness, or pain of at least a four out of ten on a ten-point scale with zero being no problem and ten being the worst possible problem) for which the patient wanted intervention. There must not have been any further planned neurotoxic chemotherapy for at least 2 months after registration.

Eligible patients were to have had a life expectancy of at least 6 months. They could not have had a previous diagnosis of diabetic or other peripheral neuropathy nor had previously used fingolimod.

At baseline, patients completed the EORTC-QLQ-CIPN20 instrument, which was then repeated weekly for four weeks.

Patients were scheduled to receive 0.5 mg of fingolimod daily for a month. As noted above, for the prevention trial, with the first dose of fingolimod patients were observed in the clinic for six hours for signs and/or symptoms of bradycardia and had a post treatment EKG at 6 hours.

Patients also completed questionnaires at baseline and weekly, to collect information regarding potential drug toxicities and compliance with taking the protocol medication. They also were questioned weekly about drug compliance and were scheduled to be called weekly by a protocol clinician, to evaluate for toxicity.

Patients enrolled on this clinical trial were treated with fingolimod 0.5 mg po qd for 4 weeks.

The tool used to evaluate CIPN was the EORTC-QLQ-CIPN20 instrument, a 20-item questionnaire, (17,18) with higher scores reflecting worse CIPN.

Adverse events were evaluated by using CTCAE criteria based on physician reporting and by patient reported outcomes determined by questionnaires.

### Statistical methods

#### Prevention trial

This was designed as a pilot study with the goal of obtaining preliminary data to support whether fingolimod would prevent CIPN in patients receiving weekly adjuvant or neoadjuvant paclitaxel therapy. The EORTC QLQ-CIPN20 questionnaire was used to measure CIPN development and persistence. The planned sample size for this trial was 20 patients. Due to only two patients being entered on this study, the data from the two patients are described/summarized in the Results Section below.

#### Treatment trial

This was designed as a pilot study to evaluate whether fingolimod, administered orally every day for 4 weeks, markedly improved symptoms in patients with established CIPN. The planned sample size was 10 patients.

The EORTC QLQ-CIPN20 questionnaire was used to measure CIPN at baseline, during treatment, and in follow up.

To assess trends in the raw CIPN sensory scores each of the 6 primary EORTC-QLQ-CIPN20 questions were plotted over time at baseline, during treatment, and in follow up. Furthermore, because each sensory symptom within the 6 primary EORTC-QLQ-CIPN20 questions appear to be important clinically, we calculated the highest (worst) numbness, tingling, and pain sensory score obtained at each timepoint. This aggregate score was plotted over time separately for each patient.

The raw scores of the Symptom Experience diary (0-10; 0= not at all, 10=as bad as can be) were used to assess toxicities related to fingolimod such as headache, fever, coughing, diarrhea, abdominal pain, lightheadedness, and nausea/vomiting.

Analyses were conducted using the SAS software version 9.4 and R software version 4.0.3.

## Results

### Prevention trial

Two patients were enrolled between 5/24/2019 and 10/7/2021. One patient did remarkably well, with minimal evidence of any neuropathy during her 12-week treatment course. However, the other patient had substantial symptoms consistent with paclitaxel-associated neuropathy. This led to her stopping paclitaxel after 8 doses.

### Treatment trial

Seven patients made up of five females and two males were registered onto the treatment of established neuropathy trial between 5/24/2019 and 10/7/2021. All of these patients had received paclitaxel. Five patients reported taking all their prescribed study doses of fingolimod, one patient stopped taking fingolimod in the second week due to a grade 1 perceived adverse eye event (temporary cloudy vision), and another patient withdrew prior to starting fingolimod (patient withdrawal).

Figure 1 illustrates the data from these six patients, with regards to the three primary symptoms, that being numbness, tingling, and pain in hands and/or feet.

**Figure 1.**
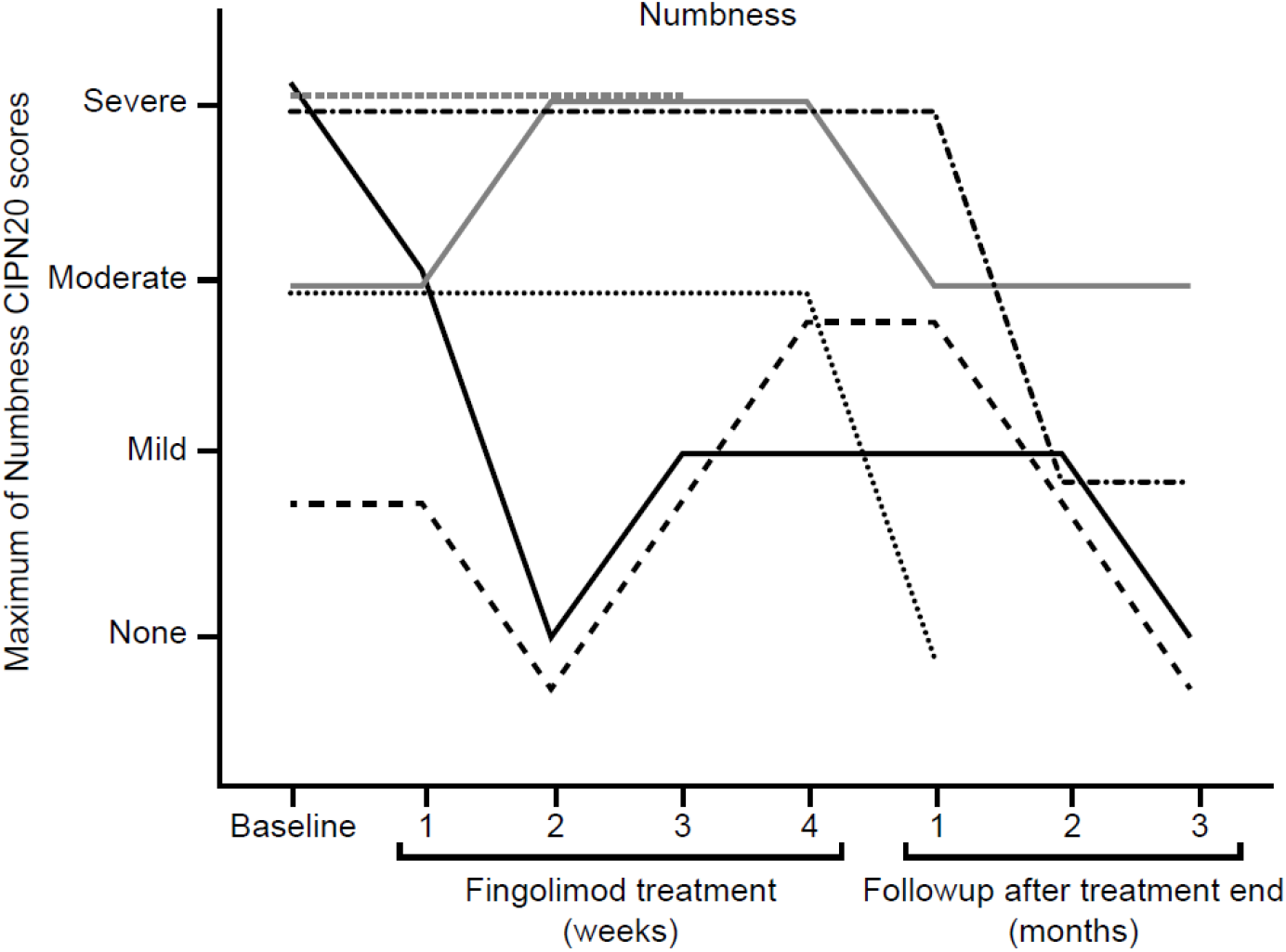
Data from 6 individual patients regarding their CIPN 20 scores for numbness (Figure 1A), tingling (Figure 1B) and pain (Figure 1C). The maximum scores (from hands and/or feet) are illustrated.

**Figure 1B.**
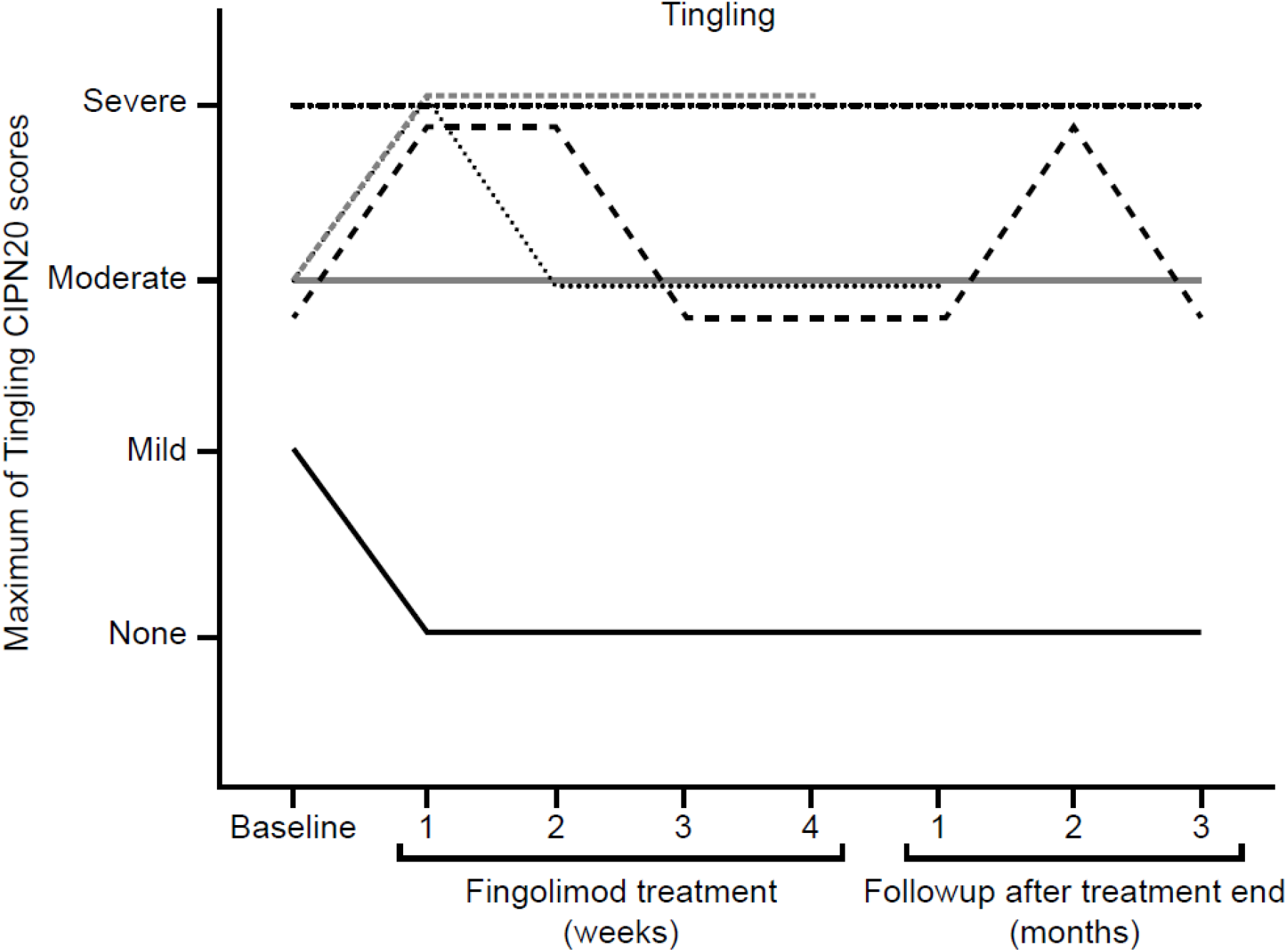

**Figure 1C.**
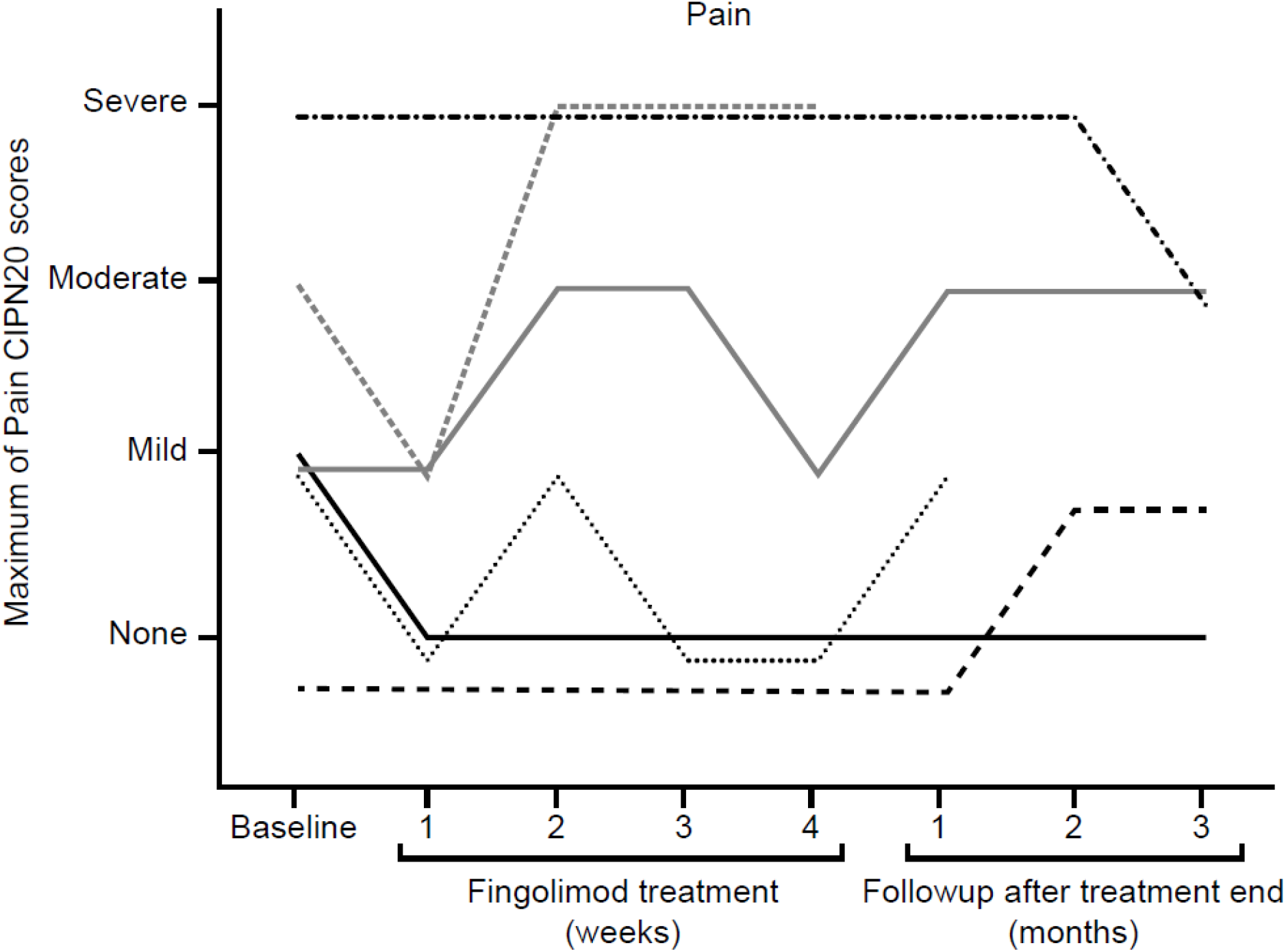
Neither the authors nor their institutions received any payments or services in the past 36 months from a third party that could be perceived to influence, or give the appearance of potentially influencing, the submitted work. This work was funded, in part, by monies from the National Cancer Institute and the Breast Cancer Research Foundation.

Regarding tolerability/toxicity of fingolimod, patients, at baseline and weekly, evaluated a number of potential fingolimod toxicities, by rating their presence on a 0-10-point scale with 0 being not at all and 10 being as bad as can be. No substantial concerns were raised regarding fever, or coughing. One single patient developed a 3-4-point worsening of lightheadedness, nausea/vomiting, abdominal pain, and diarrhea, most prominent in the second week. One additional patient had a 4-point increase in nausea/vomiting for a single week. Another patient had an 8-point increase in headache during one week while 4 others had 2-3-point increases. No patients had more than a 1-point increase in fever, or coughing. As is noted above, one patient stopped fingolimod due to an ocular complaint related to visual troubles.

## Discussion

The results of most pilot clinical trials are, as a rule, quite preliminary, by nature. Unfortunately, the results of the current two pilot clinical trials, designed to look at whether fingolimod might be able to prevent CIPN and/or treat established CIPN are even more limited as neither trial accrued the expected number of patients.

With regards to the prevention of paclitaxel neuropathy trial, one patient did quite well while the other patient did quite poorly. Thus, we cannot draw any conclusions from these two patients.

As for the treatment of established neuropathy trial, the presented figure does not reveal any good evidence that fingolimod was able to abolish established neuropathy symptoms.

While it is hard to know if the reported potential side effects were caused by fingolimod, or not, the one instance of visual trouble is of note, as fingolimod has been noted to cause macular edema. Additionally, retinal hemorrhage and retinal vein occlusion have been associated with this drug.

The complex process of monitoring patients for several hours following the initial fingolimod dose created a substantial problem of starting patients on this drug. Of note, another S1PR1 antagonist, ozanimod also decreases chemotherapy-induced neuropathy in animal models. Oxanimod has not been associated with the same temporary bradycardia that results from fingolimod and, thus, would potentially be a clinically friendlier agent.

## Data Availability

All data produced in the present work are contained in the manuscript

